# The Action Potential of an Applicant: A Seven-Year Study of Cardiac Electrophysiology Fellowship Match Outcomes (2019-2025)

**DOI:** 10.1101/2025.11.18.25340535

**Authors:** Sebastian Balda, Matias Panchana-Lascano, Ivo Diaz-Djevoich, Rodolfo Kronfle, Geovanna Minchalo-Ochoa, Thomas Leone-Berry

**Author notes:** **Corresponding Author:** Matias Panchana-Lascano, MD. Summa Veritas Medical Research. Hospital Clinica Kennedy, Office 31-A. Av. del Periodista y Callejón 11-A, N.O Kennedy. Guayaquil 090510, Ecuador. **Authors emails:**Sebastian BALDA, Ivo DIAZ-DJEVOICH, Rodolfo KRONFLE, Geovanna MINCHALO-OCHOA, Thomas LEONE-BERRY. **Funding Statement:** This research did not receive any specific grant from funding agencies in the public, commercial, or not-for-profit sectors. The study was entirely self-financed by the authors. All costs associated with the development, execution, and dissemination of this research, including materials, data collection, analysis, and manuscript preparation, were covered personally by the authors. **Authors’ Consent for Publication:** We, the authors of the manuscripts entitled “The Action Potential of an Applicant: A Seven-Year Study of Clinical Cardiac Electrophysiology Fellowship Match Outcomes” hereby declare that we have participated in the conception, design, and execution of the research work. We confirm that we have reviewed the final version of the manuscript and approved it for submission to ***Circulation: Arrhythmia and Electrophysiology***. Furthermore, we understand that this submission implies that the manuscript, in whole or in part, has not been published previously and is not under consideration for publication elsewhere. We affirm that all listed authors have agreed to be named as contributors to this work and have read and approved the final manuscript. **Permission to reproduce material from other sources:** Not applicable. **Clinical trial registration:** Not applicable. **Ethics Approval:** Not applicable. **Authorship confirmation/contribution statement: Sebastian BALDA:** Conceptualization, Methodology, Validation, Formal analysis, Supervision, Visualization, Project administration, Writing - original draft, Writing - review & editing, Investigation, Data curation. **Mathias PANCHANA-LASCANO:** Conceptualization, Methodology, Data curation, Investigation, Validation, Visualization, Writing - original draft, Writing - review & editing, Formal analysis, Project administration, Supervision. **Ivo DIAZ-DJEVOICH:** Conceptualization, Methodology, Validation, Investigation, Formal analysis, Writing - review & editing, Writing - original draft, Visualization, Data curation. **Rodolfo KRONFLE:** Conceptualization, Methodology, Validation, Investigation, Formal analysis, Writing - review & editing, Writing - original draft, Visualization, Data curation. **Geovanna MINCHALO-OCHOA:** Conceptualization, Methodology, Validation, Investigation, Formal analysis, Writing - review & editing, Writing - original draft, Visualization, Data curation. **Thomas LEONE-BERRY:** Conceptualization, Methodology, Validation, Formal analysis, Supervision, Project administration, Writing - review & editing.

## Abstract

**Background:** Clinical Cardiac Electrophysiology (CCEP) is a critical subspecialty; however, comprehensive information on fellowship match data remains limited. This study analyzes the CCEP fellowship match from 2019 to 2025 to provide insight into applicant trends, academic backgrounds, and match outcomes.

**Methods:** A retrospective cohort study was conducted using data from the National Resident Matching Program from 2019 to 2025. The analyzed variables included applicant volume, available positions and programs, match rates, and applicants’ academic backgrounds. Statistical analyses included descriptive statistics, chi-square, and Pearson correlation with p-value of <0.05 was considered statistically significant.

**Results:** Over the study period, data were collected from 980 applicants and 953 available positions, of which 867 were filled (91% match rate). The academic backgrounds of matched applicants were 48% U.S. MD, 29% Non-U.S. IMG, and 13% U.S. IMG. Most applicants (77%) matched within their top three program choices. Significant trend changes included a 99% increase in applicants (p<.001) and a 93% decrease in unfilled programs (p<.001). Unfilled positions showed strong negative correlations with being among applicants’ top three choices (r=-0.952, p<.001) and with having a higher number of U.S. MD applicants (r=-0.885, p=0.008). Matching into a first-choice program was significantly correlated with being a U.S. IMG (r=0.809, p=0.028) or a Non-U.S. IMG (r=0.871, p=0.011).

**Conclusion:** The CCEP fellowship match has increased in applicant volume and improved match rates over time. Successful matches are linked to popularity among applicants, particularly U.S. MDs, while IMGs show a positive correlation with matching into their first-choice programs.

## Introduction

Doctors following the match process to obtain a fellowship are an essential part of the healthcare system within the United States of America (USA). The organizations that coordinate the match between applicants and fellowship programs in the USA are the National Resident Matching Program (NRMP) and the Specialty Matching Service (SMS). One of these fellowships is Clinical Cardiac Electrophysiology (CCEP), which is a fundamental part of cardiology care due to the high prevalence of diseases related to electrical abnormalities in the heart^1^. In 2021, the CCEP program directors elected to go to a “all in” policy regarding applicant selection. Meaning any programs that wished to take any applicants through the match, must come from NRMP first^2^.Notwithstanding its importance in the field of medicine, there is limited data analyzing NRMP applicants matching academic backgrounds, match rates, and the competitiveness of CCEP fellowship positions.

Recent studies on fellowships of Cardiovascular disease, Pediatric Cardiology, and Advanced Heart Failure and Transplant Cardiology have demonstrated differences and challenges within each specific cardiology fellowship specialty. For instance, Pediatric Cardiology presents applicants match rates of 96,1%^3^ while Cardiovascular disease presents 66% match rates for applicants^4^. Pressing issues in other specific cardiology fellowships have been described, such as Advanced Heart Failure and Transplant Cardiology, which currently present an increasing rate of unfilled training positions in the match^5^. Other differences include rates of applicants matching into preferred choices and the academic background of the workforce of each fellowship. However, the CCEP fellowship remains underexplored, highlighting the importance of understanding the field’s tendencies.

This study aims to recognize applicant characteristics for the CCEP fellowship. Furthermore, to analyze the trends over time in match rate, unfilled positions, new positions, and applicant choice. To better inform applicants, program directors, and policymakers about subspecialty interests.

## Methods

This study is a retrospective cohort analysis of CCEP fellowship applicant data reported in the U.S. NRMP Specialty Matching Service from 2019 to 2025, designed to maintain homogeneity of the collected data. Data on the CCEP fellowship were gathered for each individual year within this period. The dataset used in this study is available as supplementary material (File 1).

The variables collected for the study included the number of applicants, available positions, programs, and positions that were filled or unfilled. Among the filled positions, applicants were categorized according to their academic backgrounds, reported as U.S. MD, U.S. DO, Canadian, U.S. IMG, and Non-U.S. IMG. In addition, data on applicants’ desired program matches were analyzed, classified as first choice, second choice, third choice, greater than third choice, and unmatched.

Descriptive analysis was conducted using numbers and percentages, while inferential analysis was performed using the chi-square test of goodness of fit to identify temporal trend changes. Pearson correlation was applied to explore associations between variables related to programs and unfilled positions, as well as applicants’ academic backgrounds and successful matches in their preferred programs. All inferential analyses were performed using SPSS version 27, and a p-value of <0.05 was considered statistically significant.

## Results

This seven-year study analyzed data from 980 applicants, including their academic backgrounds: 442 U.S. MD graduates and 538 from other academic backgrounds. The positions available for CCEP fellowship were 953, of which 867 were filled, representing an average fill rate of 91%. On the other hand, 82 positions remained unfilled.

A period of significantly increased interest in the CCEP fellowship was observed, as the number of applicants rose from 89 in 2019 to 177 in 2025—an increase of 99% (χ^2^ = 34.1, p < .001). Regarding unfilled programs, the number decreased from 45 in 2019 to just 3 in 2025, representing a 93% reduction (χ^2^ = 123, p < .001).

The number of available positions showed no significant change, increasing from 130 in 2019 to 152 in 2025—a moderate increase of 17% (χ^2^ = 3.49, p = 0.746). Similarly, the number of available programs increased from 83 in 2019 to 98 in 2025, an increase of 12% (χ^2^ = 3.67, p = 0.721). Further details are showcased in Figure 1.

**Figure 1.**
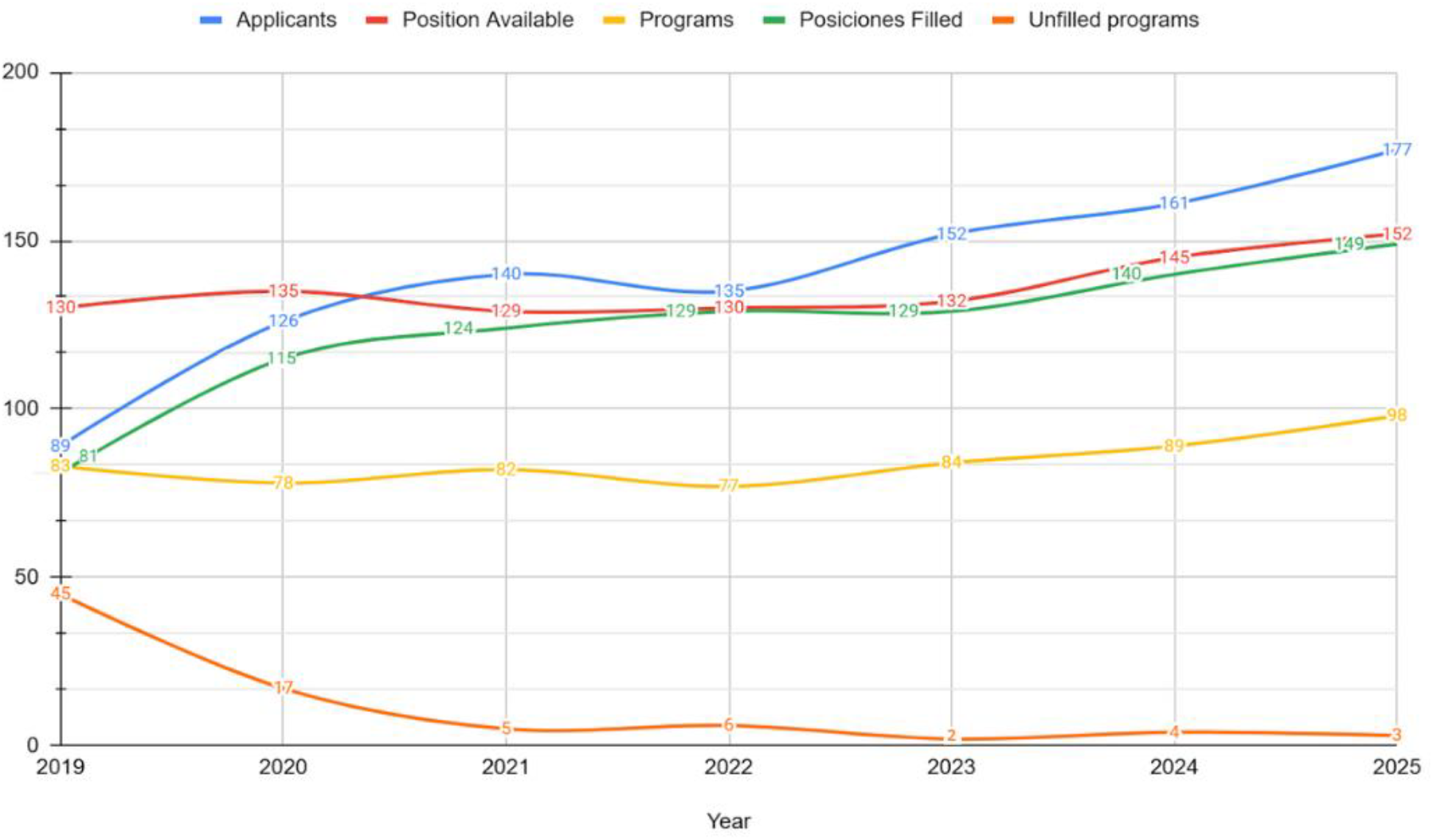
Number of applicants, positions available, programs, positions filled, and unfilled.

The most prevalent academic background among the matched applicants was U.S. MD, with 414 individuals matching, corresponding to 48% of all matches. Moreover, IMGs represented 42% of matched applicants. Non-U.S. IMGs filled 251 positions, accounting for 29% of matches, followed by U.S. IMGs with 112 filled positions, representing 13% of matches. U.S. DO graduates had 76 matches, corresponding to 9% of the total, while Canadian applicants accounted for 8 matches, representing 1% of all matches.

U.S. MD applicants rose from 39 in 2019 to 63 in 2025, an increase of 62% (χ^2^ = 14.3, p = 0.026). The number of filled positions increased from 81 in 2019 to 149 in 2025, an 84% rise (χ^2^ = 23.1, p < .001).A significant change in the match trend was observed among U.S. MD applicants, increasing from 37 in 2019 to 56 in 2025— a 51% rise (χ^2^ = 13.3, p = 0.038)—with a peak in 2024 at 73. U.S. IMGs also showed significant trend changes, increasing from 7 in 2019 to 28 in 2025, a 75% increase (χ^2^ = 15.9, p = 0.014). On the other hand, non-significant trend changes were observed among U.S. DO applicants, with matches increasing from 5 in 2019 to 14 in 2025, a 1.8% increase (χ^2^ = 8.37, p = 0.212), and among non-U.S. IMGs, from 31 in 2019 to 49 in 2025, a 58% increase (χ^2^ = 9.95, p = 0.127). The results are presented in detail in Figure 2.

**Figure 2.**
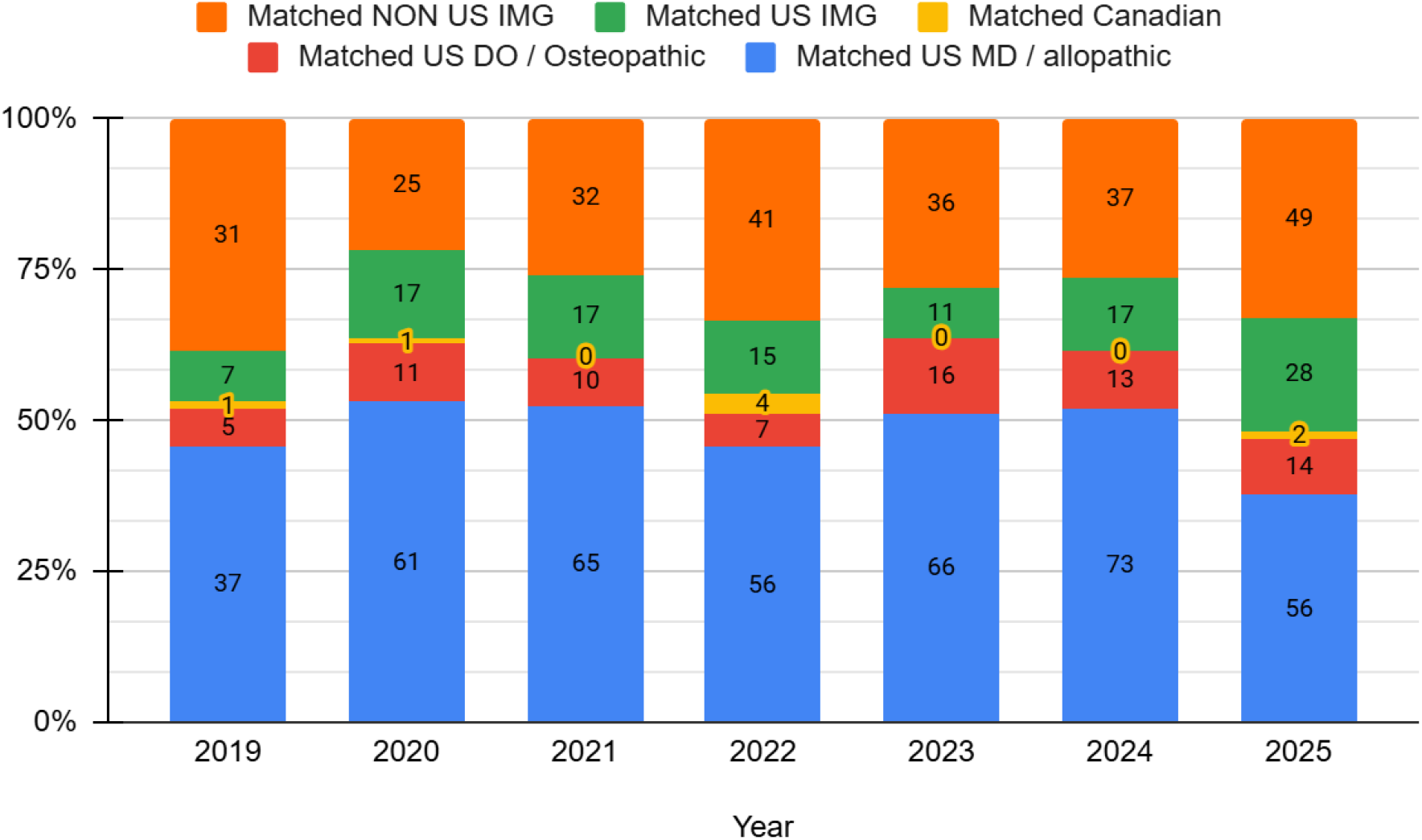
Number of matched applicants for their academic backgrounds per year with their respective percentage represented in colors.

Most matches were within applicants’ top 3 choices, with 672 (77%), from which the applicants who matched into their first choice were 496 (57%), second choice with 99 (11%), and third choice with 77 (9%). Additionally, applicants who matched on a greater-than-third choice, with 189 (22%). Finally, with 119 reported as unmatched. Detailed information is illustrated in Figure 3

**Figure 3.**
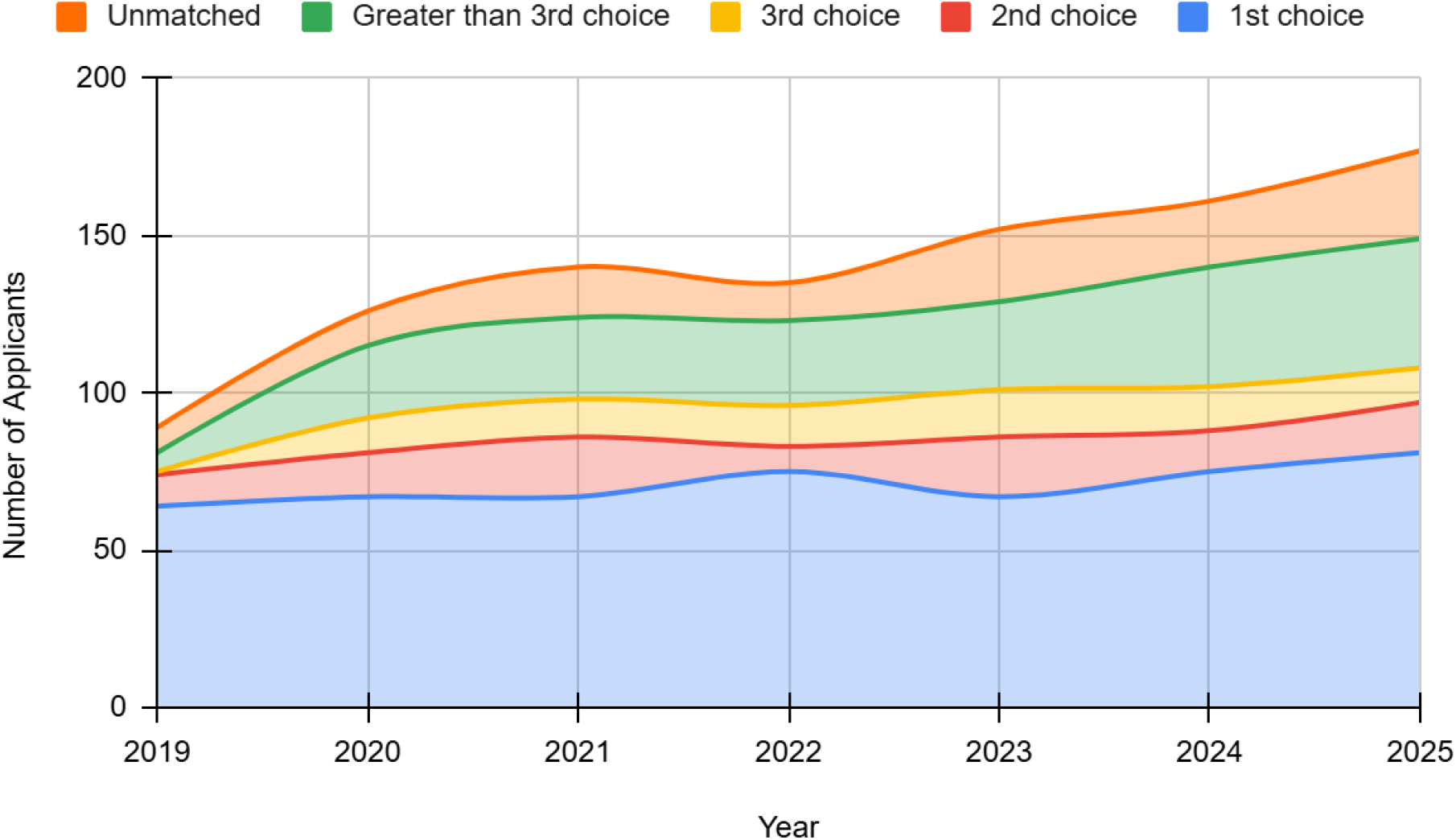
Preferred choices of Matched applicants; NRMP, National Resident Matching Program.

Significant competitiveness in CCEP fellowships was demonstrated as matches for positions ranked lower than the third choice surged from 6 (3%) in 2019 to 41 (22%) in 2025 (χ^2^ = 28.7, p < .001). Secondly, the number of unmatched applicants more than tripled, rising significantly from 8 (7%) in 2019 to 28 (24%) in 2025 (χ^2^ = 18.6, p = 0.005). This indicates growing competitiveness, with more candidates either matching farther down their rank lists or not matching at all.

In contrast, the overall match rate for applicants’ top three choices showed a non-significant upward trend, increasing from 75 (11%) in 2019 to 108 (16%) in 2025 (χ^2^ = 6.94, p = 0.327). A detailed breakdown revealed similarly non-significant fluctuations among individual top choices: first choice matches increased from 64 (79%) in 2019 to 81 (81%) in 2025 (χ^2^ = 3.23, p = 0.779); second choice rose from 10 (12%) in 2019 to 16(11%) in 2025 (χ^2^ = 7.56, p = 0.273); and the third choice matches increased from 1 (1%) in 2019 to 11 (7%) in2025 (χ^2^ = 11.8, p = 0.066).

### Unfilled Programs correlations with matched applicants’ academic backgrounds

The analysis demonstrated a significant correlation between unfilled programs and being within the applicants’ top three choices (r = −0.952, p < .001). The strongest academic background associated with fewer unfilled programs was matched U.S. MD applicants (r = −0.840, p = 0.018). Additional significant associations were observed with the number of applicants (r = −0.893, p = 0.007), particularly the number of U.S. MD applicants (r = −0.885, p = 0.008).

Other academic backgrounds did not show significant correlations with unfilled programs: U.S. DO (R = −0.719, p = 0.069), U.S. IMG (R = −0.593, p = 0.161), Non-U.S. IMG (R = −0.484, p = 0.271), and Canadian applicants (R = −0.013, p = 0.979).

### Applicants matching their preferred programs and their academic backgrounds

Our study showed a significant association between applicants matching into their first-choice programs and foreign medical school backgrounds. Matched U.S. IMGs (r = 0.809, p = 0.028) and matched Non-U.S. IMGs (r = 0.871, p = 0.011) demonstrated strong, statistically significant correlations. In contrast, other groups did not show significant associations, including matched U.S. MDs (r = 0.255, p = 0.581), matched U.S. DOs (r = 0.332, p = 0.467), and matched Canadian applicants (r = 0.482, p = 0.273).

## Discussion

The fast expansion in applicant volume in this study resembles national growth among other cardiology subspecialties but exceeds the relative increase seen in most^6^. The NRMP reports that the number of General Cardiovascular Disease fellowship applicants increased by 15% between 2019 and 2023, while the numbers for interventional cardiology and advanced heart failure/transplant cardiology grew by 20% and 18%, respectively, in 2024^7^. A great increase of nearly double in CCEP applicants in the same timeframe may indicate the increased interest in new technologies like 3D mapping of the heart’s electrical system, catheter procedures, and modern pacemaker devices, which may have made the specialty more attractive to trainees. Such advances have expanded the number of procedures done per year^8^.

Across the whole array of medical subspecialties, the trends in unfilled positions for fellowships vary widely. According to the NRMP and their SMS data from 2019 to 2025, most internal medicine fellowships grew in programs, positions, and applicants. However, several reports show persistence of unfilled rates in certain fields. Advanced Heart Failure and Transplant Cardiology (AHFTC) has notably emerged among the cardiovascular fellowships with the most difficult slots to fill. An ACC commentary noted that in one match cycle, of 127 positions offered, only 61 (48%) were filled, and just 21 of 75 programs (28%) succeeded in filling all of their slots^9^. Recent cross-sectional analysis covering 2020-2025 documented that the overall fill rate across many years was only 58%, and unfilled positions rose from 30% to 52% over that interval^5^.These findings show a gap between training demand and applicant supply in AHFTC. Several surveys from cardiology professionals highlight some discouraging factors, including demanding clinical workload, extended hours, and perceived lower compensation relative to other cardiology fellowships^10^.

In contrast, CCEP demonstrated a more favorable trend. Despite having just a 17% increase from 2019 to 2025 in available fellowship positions, there was a 93% reduction in unfilled programs, dropping from 45 unfilled positions to 3 in the study timeframe. During this same period, filled positions rose by 84% and applicant volume nearly doubled; which reflects rising competitiveness and better matching between applicant demand and program availability. Aligning with previous analysis describing growth in the annual number of applicants and decrease in unfilled positions^11^. When performing statistical analysis on successful matches and its relation to the other variables in the study, the “program popularity”, which was based on applicants’ top 3 ranking, was statistically significant. In addition, programs with a higher proportion of U.S MD applicants showed statistical significance in regards to decreasing unfilled positions. While this does not establish causality, it may suggest that programs perceived as more competitive tend to attract applicants with greater choice and, in turn, are more likely to fill their slots.

Our study results show consistent trends in the applicant’s pool for CCEP among the cardiology-related fellowships. For most of the match cycles, the majority of matched applicants consist of U.S. MDs, followed by Non-U.S. IMGs, U.S. IMGs, and U.S. DO, particularly in last year’s match cycle having 37.58%, 32.89%, 18.79%, 9.4%, and 1.34% respectively. However for the fellowship of cardiovascular disease match rates per applicant type between 2010 and 2021 were majoritarily achieved by U.S. MDs ranging between 83% and 93%; with the highest 93% in 2016 and the lowest 83% both in 2010 and 2021. The second distinction is the undisclosed non-U.S. MDs ranging between 41% and 62% match rate with the highest in 2016 and the lowest in 2010^4^. Regarding the fellowship of adult congenital heart disease, it is a relatively novel training program, with its recognition as a distinct subspeciality dating to 2011^12^. As of the time of this study, there has been no published analysis of applicants, their demographics, or position availability trends.

The results of this study indicate the number of applicants for the CCEP fellowship almost doubled, yet the proportion of non-U.S. IMGs remained consistent at 42% of matched trainees, highlighting their sustained contribution in the subspecialty. Moreover, the 40% participation of non-U.S. IMGs showcases the global appeal of this subspecialty^13^.The variation of applicant type provides insight into the evolving structure of the workforce^13,14^.

CCEP plays a critical role in the intervention and management of cardiac arrhythmias. Modern techniques like catheter ablation, electroanatomic mapping, and pacemaker devices have drastically changed patients’ outcomes, reducing hospitalizations and sudden cardiac deaths^15^. The increasing demand for these procedures has been accompanied by the parallel increase in applicants and well-trained electrophysiology workforce. In this context, the findings of our study have provided insight into the evolving dynamics of the CCEP fellowship workforce pathway.

## Conclusion

The CCEP fellowship match has seen an increase in applicant volume and improved fill rates. Program match success is closely tied to popularity among applicants, particularly U.S. MDs, while International Medical Graduates show a distinct correlation with matching at their first-choice program

## Data Availability

Data extracted from the National Resident Matching Program (NRMP) and the Specialty Matching Service (SMS) supporting this work is available as csv supplementary file 1.

